# Effects of Continuous Oral Intake of DR.ERGO^®^ Ergothioneine Capsules on Skin Status: A Randomized, Double-Blind, Placebo-Controlled Trial

**DOI:** 10.1101/2025.10.16.25337962

**Authors:** Xiang Ma, Adachi Masaru, Wei CUI, Jinhui Du, Huimin Liu

## Abstract

**Background:** Ergothioneine (ergothioneine) is a naturally occurring antioxidant with emerging evidence supporting its role in cellular protection and anti-aging. However, clinical data on its cosmetic benefits remain limited.

**Objective:** To evaluate the effects of daily oral supplementation with DR.ERGO^®^ ergothioneine capsules on skin condition in healthy women reporting signs of skin aging.

**Methods:** In this 8-week, randomized, double-blind, placebo-controlled trial, 66 women aged 35–59 years were assigned to receive either DR.ERGO^®^ ergothioneine (30 mg/day) or placebo. Skin parameters including melanin index, erythema index, glossiness, elasticity (R2, R5, R7), spot count, wrinkle count were evaluated at baseline, 4 weeks, and 8 weeks.

**Results:** Compared to placebo, the DR.ERGO^®^ ergothioneine group showed significantly greater improvements in melanin and erythema reduction, skin glossiness, elasticity, and wrinkle and spot reduction (all p < 0.01). No adverse events were reported.

**Conclusion:** Continuous oral intake of DR.ERGO^®^ ergothioneine significantly improved skin aging parameters, with excellent safety and tolerability. These findings suggest DR.ERGO^®^ ergothioneine may serve as an effective oral anti-aging agent.

## 1. Introduction

Skin aging is a complex biological process driven by both intrinsic and extrinsic factors, including chronological aging, oxidative stress, ultraviolet (UV) radiation, and environmental pollution (Zhang & Duan, 2018). These factors lead to structural and functional changes in the skin, such as increased pigmentation, loss of elasticity, dullness, and the formation of wrinkles and spots (Shin et al., 2019). In recent years, the growing consumer demand for skin health and anti-aging solutions has spurred interest in nutritional approaches—particularly oral supplementation with bioactive compounds known as “nutricosmetics” (Rahman et al., 2024).

Ergothioneine (ergothioneine) is a naturally occurring thiol compound classified as a cytoprotective antioxidant (Borodina et al., 2020). First discovered in ergot fungi and later identified in various food sources such as mushrooms and oat bran, ergothioneine is unique in its high affinity for mitochondrial accumulation and selective uptake via the organic cation transporter OCTN1 (Tucker et al., 2019). Unlike classical antioxidants, ergothioneine resists auto-oxidation, chelates reactive oxygen species (ROS), and protects biomolecules such as DNA and proteins from oxidative damage (Fu & Shen, 2022). Preclinical studies have demonstrated its potential in attenuating oxidative stress, inflammation, and mitochondrial dysfunction—pathways central to the skin aging process (Apparoo et al., 2022; Hanayama et al., 2024).

Despite these promising properties, there is a paucity of human clinical data regarding the effects of ergothioneine on dermatological outcomes. Most studies to date have focused on its systemic antioxidant and neuroprotective roles, with only limited evidence available on its efficacy in improving visible signs of skin aging (Chunyue Zhang, 2023; Hanayama et al., 2024). Given its targeted accumulation in mitochondria—critical hubs of oxidative metabolism—and potent antioxidant properties, ergothioneine is hypothesized to modulate redox-sensitive pathways involved in cellular aging. These characteristics suggest its potential as an oral agent for mitigating skin aging. However, clinical validation using rigorous methodology is urgently needed.

To address this gap, we conducted an 8-week, randomized, double-blind, placebo-controlled clinical trial to evaluate the effects of daily oral supplementation with 30 mg of ergothioneine (DR.ERGO^®^) on skin parameters in healthy adult women aged 35–59 years who reported subjective signs of skin aging. Objective measurements—including melanin and erythema indices, skin glossiness, elasticity, and wrinkle and pigmentation counts—were used to comprehensively evaluate changes in skin condition.

This study aims to provide clinical evidence for the efficacy and safety of ergothioneine as an oral nutricosmetic ingredient. By integrating instrumental assessments with participant-reported outcomes, we sought to capture both the physiological and perceptual benefits of ergothioneine supplementation. Our findings contribute to the growing body of research on nutritional strategies for promoting skin health and delaying the onset of visible skin aging.

## 2. Materials and methods

### 2.1. Subjects and Methods

#### 2.1.1. Study Design and Ethical Review

This randomized, double-blind, placebo-controlled trial was conducted by the Japan Clinical Trial Association (JACTA), under the supervision of the principal investigator. All measurements were performed in JACTA’s internal laboratory.

The trial was prospectively registered on the UMIN Clinical Trials Registry (UMIN000055792) and strictly adhered to the Declaration of Helsinki (2013 revision) and the Ethical Guidelines for Life Science and Medical Research (implemented March 2021).

Approval was obtained from the Pharmaceutical Affairs Law Ethical Review Board. Prior to enrollment, all participants received detailed written and oral explanations regarding the trial’s objectives and procedures. Written informed consent was obtained from each participant before initiation of the study.

#### 2.1.2. Subjects

JACTA will be accepting applications from the public through Breakthrough Co., Ltd. (Tokyo). Eligible subjects were healthy volunteers who met all inclusion criteria, none of the exclusion criteria, and voluntarily agreed to consume the test product.

##### 2.1.2.1. Inclusion Criteria

1. Healthy female subjects aged 35–59 years;
2. Individuals self-reporting perceived skin deterioration (e.g., loss of radiance, dullness, spots, etc.).

##### 2.1.2.2. Exclusion Criteria

1. History of food allergies;
2. Pregnant or lactating women;
3. Facial inflammation or open wounds;
4. Current use of medications that may interfere with trial results;
5. Regular intake of health supplements that may affect trial outcomes;
6. Under medical treatment or holding an active physician’s prescription;
7. Ongoing hormone therapy;
8. Semi-permanent makeup (e.g., eyelash extensions, eyeliner tattooing) around the eyes;
9. Participation in another clinical trial within the past month or planned participation during this study;
10. Deemed ineligible by the principal investigator.

#### 2.1.3. Sample Size

Based on preliminary data, a sample size of 60 subjects was determined to achieve 80% statistical power at a 5% significance level, accounting for potential dropouts.

#### 2.1.4. Test Products

The test product, DR.ERGO^®^ Ergothioneine capsules and placebo capsules, were provided by Shanghai Ergothioneine Biotechnology Group Co., Ltd. Participants were instructed to take one capsule daily for 8 weeks, consumed with warm water before breakfast.

To ensure blinding, the test product (ergothioneine) and placebo control were indistinguishable in shape, color, and taste, and were distributed using coded labels.

**Table 1-1.**
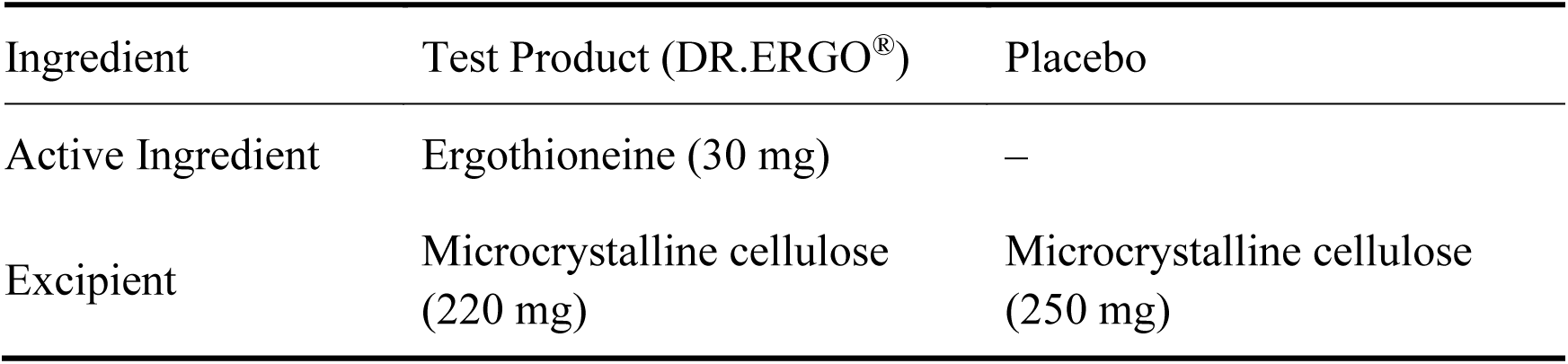
Composition of Test and Control Products.

**Table 1-2.** Nutritional Composition of Ergothioneine (per 100 g)

| Component | Amount |
| --- | --- |
| Energy | 0 kJ |
| Protein | 0 g |
| Fat | 0 g |
| Carbohydrates | 0 g |
| Sodium (Na) | 0 mg |

#### 2.1.5. Randomization

A total of 70 eligible subjects who met the inclusion criteria and none of the exclusion criteria were enrolled in the study. Random allocation was performed by an independent allocation officer not involved in the trial to minimize bias. To ensure balanced group assignment, stratification was applied based on age and BMI, dividing the 70 participants into two groups (A and B) with 35 subjects each.

The allocation details were strictly maintained in confidentiality by the allocation officer and were only disclosed to the trial investigators after database lock (unblinding). Group A received DR.ERGO^®^ Ergothioneine Capsules once daily for 8 weeks. Group B received a matched placebo under the same regimen.

#### 2.1.6. Study Schedule

The trial was conducted from October to December 2024. Evaluations were performed at three time points: baseline (pre-intervention), week 4, and week 8. Standardized measurement protocols were implemented to ensure consistency in posture, positioning, and timing of assessments across all visits.

During the study period, participants were instructed to refrain from initiating any new medications or dietary supplements that might affect the test areas; maintain their usual lifestyle patterns and complete daily logs to document compliance with study requirements. The detailed study schedule is presented in Table 2.

**Table 2.** Study Schedule.

| Phase | Consent | Intervention Period |  |  |
| --- | --- | --- | --- | --- |
|  |  | Baseline | Week 4 | Week 8 |
| Screening & Randomization | ● |  |  |  |
| Measurements |  | ● | ● | ● |
| DR.ERGO® or Placebo intake |  | √ | √ | √ |
| Daily log entries |  | √ | √ | √ |

●: Performed on measurement days √: Performed daily throughout the period

#### 2.1.7. Participant Compliance Requirements

All participants were instructed to maintain their normal lifestyle throughout the study period while adhering to the following requirements:

1. Consistency in Lifestyle Habits: maintain pre-trial dietary patterns, exercise routines, alcohol consumption, smoking habits, and sleep duration without modification;
2. Avoidance of extreme behaviors: refrain from excessive exercise, avoid sleep deprivation, prevent overeating (including banquet participation, buffet meals, or all-you-can-eat occasions) and abstain from weight loss programs;
3. Restrictions on skin-related products: prohibited use of any pharmaceuticals, quasi-drugs, or health foods claiming skin-related benefits and forbidden to receive any cosmetic treatments or therapies;
4. Medication controls: medication use was prohibited except in unavoidable circumstances, required documentation of any necessary medication (name and dosage) in daily logs;
5. Consistency in supplement use: maintain existing quasi-drug and health food regimens (dosage, frequency, and administration method), prohibited initiation of any new quasi-drugs or health supplements;
6. Pre-measurement restrictions (72-hour period): no all-nighters or sleep deprivation and avoidance of strenuous exercise (e.g., exhaustive running, swimming, mountain climbing);
7. 24-hour pre-measurement requirements: complete alcohol abstinence, ensure adequate sleep and maintain optimal physical condition.

### 2.2. Evaluation Items and Methods

At each of the four observation time points, participants were required to cleanse their skin using commercially available makeup remover and facial cleanser, and acclimate for 20 minutes in a controlled environment (temperature: 21±1°C, humidity: 50±5% RH) before measurements. VISIA measurements were performed bilaterally on both the left and right sides of the face. All other instrumental assessments were conducted at a standardized anatomical point, defined as the intersection of a vertical line descending from the outer canthus and a horizontal line extending from the alar base (nostrils). Reported values included individual measurements (left and right sides) and mean values (average of left and right).

#### 2.2.1. Melanin and Erythema Indices

Melanin and erythema levels were quantitatively assessed using the Mexameter^®^ MX 18 (Courage+Khazaka electronic GmbH) at standardized facial sites. Measurements were recorded in arbitrary units, with decreasing values corresponding to reduced pigmentation and erythema severity.

#### 2.2.2. Skin Glossiness

Skin surface reflectance was measured with the Glossymeter GL200MP (Courage+Khazaka), which incorporates optical compensation algorithms to minimize interference from skin texture and chromatic variation. Results were expressed in gloss units (GU), where higher values indicate superior light reflectance properties.

#### 2.2.3. Skin Elasticity

Biomechanical properties were evaluated using the Cutometer^®^ MPA580 (Courage+Khazaka), which quantifies three key elasticity parameters: R2 (gross elasticity), R5 (net elasticity), and R7 (biological elasticity). All measurements were normalized to a 0-1 scale, with values approaching 1.0 representing optimal elastic function.

#### 2.2.4. Pigmentation and Wrinkles

The VISIA^®^ Evolution system (Canfield Scientific) provided multidimensional assessment of hyperpigmentation and rhytides through standardized metrics including absolute spot counts, clinical severity scores, and population-based percentile rankings. Improved skin conditions were reflected by decreased spot counts and elevated percentile rankings relative to age-matched norms.

#### 2.2.5. Safety Evaluation

Safety monitoring incorporated systematic review of participant-maintained lifestyle logs and structured adverse event reporting, with particular attention to protocol compliance and product tolerability throughout the 8-week intervention period.

#### 2.2.6. Statistical processing

##### 2.2.6.1. Evaluation method

The full analysis set (FAS) was used for all statistical analyses. Continuous variables were expressed as mean ± standard deviation. Within-group comparisons versus baseline were performed using paired t-tests, while between-group comparisons and analysis of demographic variables utilized Student’s t-tests. No adjustments were made for multiple comparisons, and complete case analysis was applied given the absence of missing data. All tests were two-tailed, with statistical significance set at p<0.05. Analyses were conducted using Statcel 4 software.

##### 2.2.6.2. Change Rate Calculation Formulas

1. Between-Group Comparison (DR.ERGO^®^ vs. Placebo) 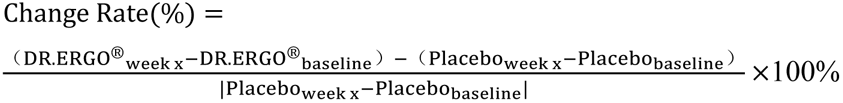
2. Within DR.ERGO^®^ Group 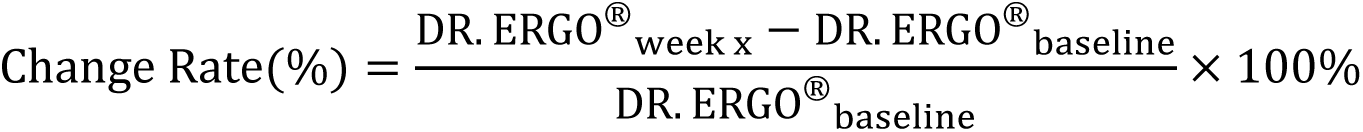
3. Within Placebo Group 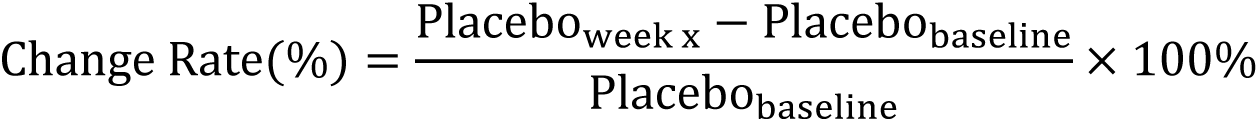

## 3. Results

### 3.1. Subject Enrollment and Demographics

The study initially enrolled 70 eligible participants, of whom 66 completed the full trial protocol (completion rate: 94.3%). Four participants discontinued intervention due to personal reasons unrelated to the study procedures. The final analysis cohort consisted of 66 female subjects aged 35-59 years, with a mean age of 44.4 ± 6.3 years, representing the target demographic for skin aging evaluation. Figure 1 presents the complete participant flow diagram following CONSORT guidelines, while Table 3 provides comprehensive baseline demographic and clinical characteristics of both intervention groups. The high retention rate and balanced baseline parameters between groups support the robustness of subsequent efficacy analyses.

**Figure 1.**
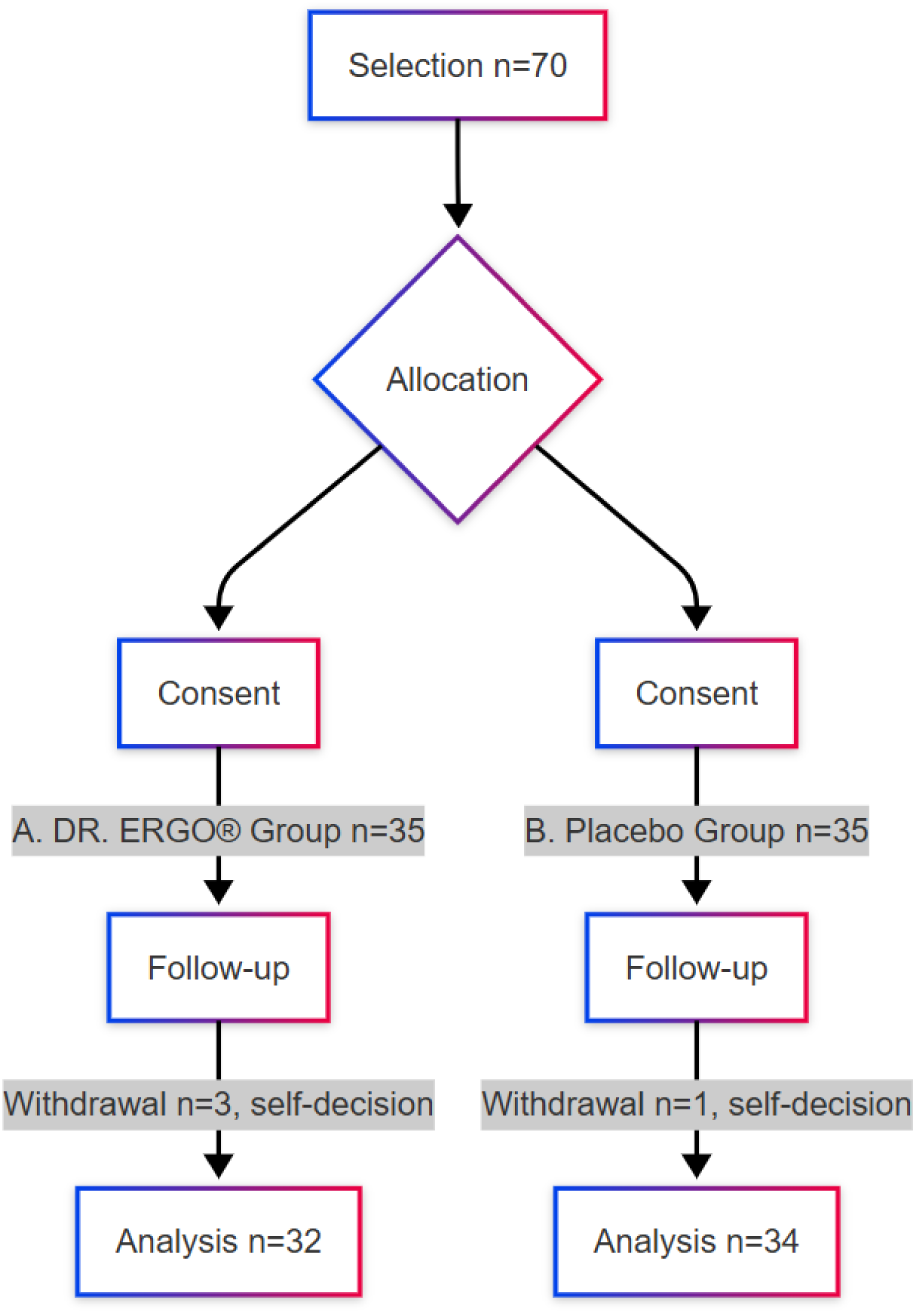
Process of Determining Subjects for Analysis.

**Table 3.** Background of Subjects Included in the Efficacy Analysis.

| Item | Unit | DR.ERGO® Group (n=32) | Control Group (n=34) |
| --- | --- | --- | --- |
| Age * | Years | 44.6 ± 6.4 | 44.9 ± 6.6 |
| BMI * | kg/m<sup>2</sup> | 20.8 ± 3.1 | 20.7 ± 2.1 |
| Menopause * | Index | 128.08 ± 28.79 | 131.57 ± 36.56 |
| Erythema * | Index | 254.36 ± 48.13 | 242.78 ± 45.28 |
| Scale * | Index | 4.85 ± 1.33 | 5.44 ± 1.41 |
| Elasticity R2 * | Unitless | 0.69 ± 0.07 | 0.67 ± 0.07 |
| Elasticity R5 * | Unitless | 0.48 ± 0.06 | 0.46 ± 0.08 |
| Elasticity R7 * | Unitless | 0.34 ± 0.04 | 0.32 ± 0.04 |

Values are expressed as mean ± standard deviation.
\* No significant difference between groups.

### 3.2. Melanin and Erythema Indices

The quantitative changes are shown in Table 4. Compared to placebo, the DR.ERGO^®^ group showed significantly greater reductions in both melanin (136.7% and 202.9% at weeks 4 and 8, respectively) and erythema (169.8% and 215.1%) (all p<0.01).

**Table 4.** Longitudinal Changes in Melanin, Erythema, Glossiness, and Elasticity Parameters.

| Item<br>(Unit) | Time Point | Measured values |  | p-value<sup>2)</sup> |
| --- | --- | --- | --- | --- |
|  |  | DR.ERGO<sup>®</sup><br>Group (n=32)<sup>1)</sup> | Placebo Group<br>(n=34)<sup>1)</sup> |  |
| Melanin<br>(Index) | a Before start | 128.1 ± 28.8 | 131.6 ± 36.6 |  |
|  | b After 4 weeks | 126.4 ± 28.3 | 136.1 ± 35.5 |  |
|  | Δa-b | -1.7 ± 3.7* | 4.6 ± 3.9** | <0.001<sup>##</sup> |
|  | c After 8 weeks | 122.3 ± 28.3 | 137.2 ± 36.7 |  |
|  | Δa-c | -5.8 ± 6.3** | 5.6 ± 5.0** | <0.001<sup>##</sup> |
| Erythema<br>(Index) | a Before start | 254.36 ± 48.13 | 242.78 ± 45.28 |  |
|  | b After 4 weeks | 248.45 ± 47.47 | 251.24 ± 44.15 |  |
|  | Δa-b | -5.91 ± 8.72** | 8.46 ± 5.67** | <0.001<sup>##</sup> |
|  | c After 8 weeks | 240.59 ± 45.90 | 254.74 ± 46.08 |  |
|  | Δa-c | -13.77 ± 12.94** | 11.96 ± 8.18** | <0.001<sup>##</sup> |
| Gloss<br>(Index) | a Before start | 4.85 ± 1.33 | 5.44 ± 1.41 |  |
|  | b After 4 weeks | 4.99 ± 1.35 | 5.39 ± 1.39 |  |
|  | Δa-b | 0.15 ± 0.15** | -0.06 ± 0.11** | <0.001<sup>##</sup> |
|  | c After 8 weeks | 5.31 ± 1.41 | 4.92 ± 1.19 |  |
|  | Δa-c | 0.46 ± 0.46** | -0.52 ± 0.59** | <0.001<sup>##</sup> |
| Elasticity<br>R2 | a Before start | 0.69 ± 0.07 | 0.67 ± 0.07 |  |
|  | b After 4 weeks | 0.70 ± 0.07 | 0.65 ± 0.07 |  |
|  | Δa-b | 0.01 ± 0.01** | -0.01 ± 0.03** | <0.001<sup>##</sup> |
|  | c After 8 weeks | 0.73 ± 0.06 | 0.64 ± 0.07 |  |
|  | Δa-c | 0.03 ± 0.03** | -0.03 ± 0.03** | <0.001<sup>##</sup> |
|  |  | DR.ERGO<sup>®</sup><br>Group (n=32)<sup>1)</sup> | Placebo Group<br>(n=34)<sup>1)</sup> |  |
| Elasticity<br>R5 | a Before start | 0.48 ± 0.06 | 0.46 ± 0.08 |  |
|  | b After 4 weeks | 0.49 ± 0.06 | 0.45 ± 0.08 |  |
|  | Δa-b | 0.01 ± 0.01<sup>**</sup> | -0.01 ± 0.03<sup>*</sup> | 0.001<sup>##</sup> |
|  | c After 8 weeks | 0.51 ± 0.06 | 0.43 ± 0.07 |  |
|  | Δa-c | 0.02 ± 0.02<sup>**</sup> | -0.03 ± 0.04<sup>**</sup> | <0.001<sup>##</sup> |
| Elasticity<br>R7 | a Before start | 0.34 ± 0.04 | 0.32 ± 0.04 |  |
|  | b After 4 weeks | 0.35 ± 0.04 | 0.31 ± 0.04 |  |
|  | Δa-b | 0.01 ± 0.01<sup>**</sup> | -0.01 ± 0.02<sup>**</sup> | <0.001<sup>##</sup> |
|  | c After 8 weeks | 0.37 ± 0.03 | 0.31 ± 0.04 |  |
|  | Δa-c | 0.03 ± 0.02<sup>**</sup> | -0.02 ± 0.03<sup>**</sup> | <0.001<sup>##</sup> |

Values are expressed as mean ± standard deviation.
1) \*p<0.05 and \*\*p<0.01 indicate significant differences from baseline;
2) ##p<0.01 indicates significant difference from placebo group.

Compared to baseline, DR.ERGO^®^ significantly decreased melanin (1.3% and 4.5%) and erythema (2.3% and 5.4%) at weeks 4 and 8 (all p<0.05), while placebo showed significant increases in both parameters (melanin: 3.5% and 4.3%; erythema: 3.5% and 4.9%, p<0.05).

### 3.3. Skin Glossiness Measurements

The changes in skin glossiness are presented in Table 4. Compared to placebo, the DR.ERGO^®^ group demonstrated significantly greater increases in glossiness (364.9% and 188.2% at weeks 4 and 8, respectively; all p<0.01).

Longitudinal analysis revealed that DR.ERGO^®^ significantly increased glossiness from baseline (3.0% and 9.5% at weeks 4 and 8, p<0.05), while placebo showed significant reductions (1.0% and 9.6%, p<0.05).

### 3.4. Biomechanical Elasticity Parameters

The quantitative changes in skin elasticity parameters (R2, R5, R7) are summarized in Table 4. Compared to placebo, the DR.ERGO^®^ group showed significantly greater improvements in all elasticity indices at both timepoints (R2: 183.9% and 210.5%; R5: 155.0% and 195.0%; R7: 215.0% and 252.6% at weeks 4 and 8, respectively; all p<0.01).

Compared to baseline, DR.ERGO^®^ significantly increased R2 (1.7% and 4.7%), R5 (1.5% and 4.9%), and R7 (3.7% and 8.4%) at weeks 4 and 8 (all p<0.05). In contrast, placebo significantly decreased all parameters (R2: −2.1% and −4.4%; R5: −2.8% and −5.5%; R7: −3.3% and −5.7%; all p<0.05).

### 3.5. Results of Spots, Latent Spots, Brown Spots, and Wrinkles

#### 3.5.1. Counts

The changes in quantitative measures are presented in Table 5-1. Compared to placebo, the DR.ERGO^®^ group showed significantly greater reductions in all measured counts at both timepoints: visible spots (251.2% and 275.2% at weeks 4 and 8, respectively), latent spots (185.5% and 187.9%), brown spots (203.1% and 204.6%), and wrinkles (139.6% and 152.9%) (all p<0.01).

**Table 5-1.**
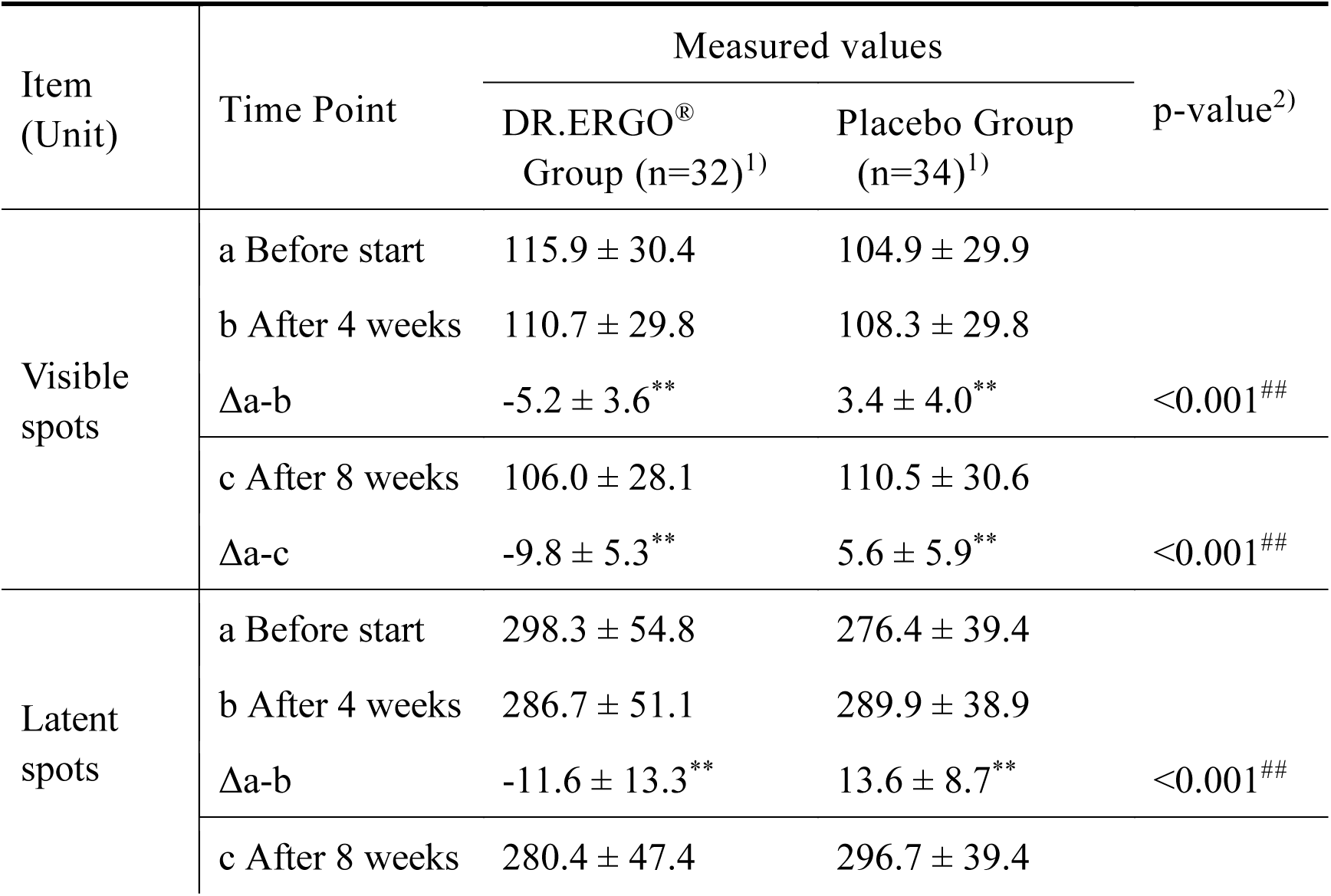
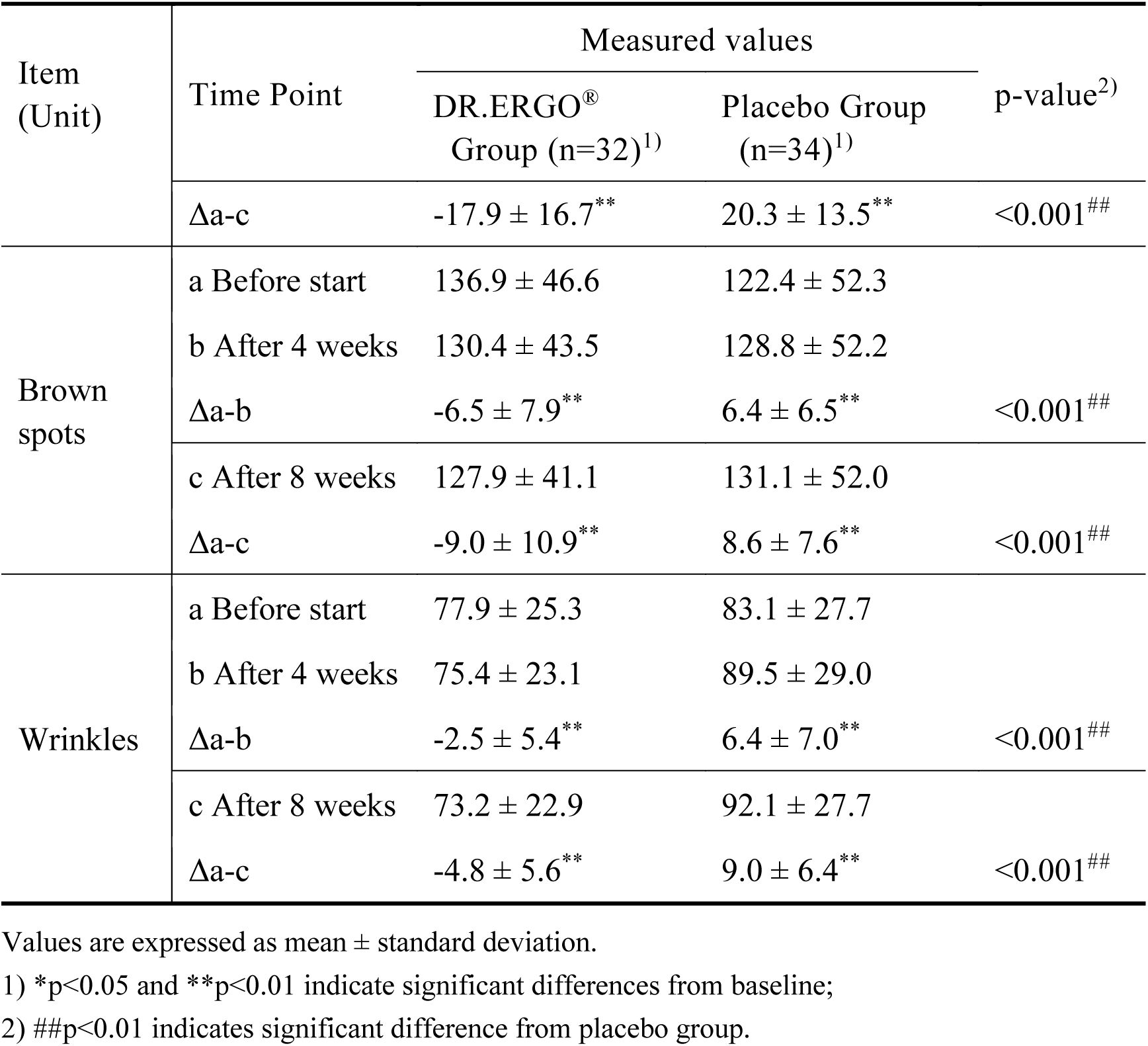
Longitudinal Changes in Spot Counts (Visible Spots, UV Spots, Brown Spots, and Wrinkles)

| Item<br>(Unit) | Time Point | Measured values |  | p-value<sup>2)</sup> |
| --- | --- | --- | --- | --- |
|  |  | DR.ERGO<sup>®</sup><br>Group (n=32)<sup>1)</sup> | Placebo Group<br>(n=34)<sup>1)</sup> |  |
| Visible spots | a Before start | 115.9 ± 30.4 | 104.9 ± 29.9 |  |
|  | b After 4 weeks | 110.7 ± 29.8 | 108.3 ± 29.8 |  |
|  | Δa-b | -5.2 ± 3.6** | 3.4 ± 4.0** | <0.001## |
|  | c After 8 weeks | 106.0 ± 28.1 | 110.5 ± 30.6 |  |
|  | Δa-c | -9.8 ± 5.3** | 5.6 ± 5.9** | <0.001## |
| Latent spots | a Before start | 298.3 ± 54.8 | 276.4 ± 39.4 |  |
|  | b After 4 weeks | 286.7 ± 51.1 | 289.9 ± 38.9 |  |
|  | Δa-b | -11.6 ± 13.3** | 13.6 ± 8.7** | <0.001## |
|  | c After 8 weeks | 280.4 ± 47.4 | 296.7 ± 39.4 |  |
|  |  | DR.ERGO<sup>®</sup><br>Group (n=32)<sup>1)</sup> | Placebo Group<br>(n=34)<sup>1)</sup> |  |
|  | Δa-c | -17.9 ± 16.7** | 20.3 ± 13.5** | <0.001<sup>##</sup> |
| Brown spots | a Before start | 136.9 ± 46.6 | 122.4 ± 52.3 |  |
|  | b After 4 weeks | 130.4 ± 43.5 | 128.8 ± 52.2 |  |
|  | Δa-b | -6.5 ± 7.9** | 6.4 ± 6.5** | <0.001<sup>##</sup> |
|  | c After 8 weeks | 127.9 ± 41.1 | 131.1 ± 52.0 |  |
|  | Δa-c | -9.0 ± 10.9** | 8.6 ± 7.6** | <0.001<sup>##</sup> |
| Wrinkles | a Before start | 77.9 ± 25.3 | 83.1 ± 27.7 |  |
|  | b After 4 weeks | 75.4 ± 23.1 | 89.5 ± 29.0 |  |
|  | Δa-b | -2.5 ± 5.4** | 6.4 ± 7.0** | <0.001<sup>##</sup> |
|  | c After 8 weeks | 73.2 ± 22.9 | 92.1 ± 27.7 |  |
|  | Δa-c | -4.8 ± 5.6** | 9.0 ± 6.4** | <0.001<sup>##</sup> |

Values are expressed as mean ± standard deviation.
1) \*p<0.05 and \*\*p<0.01 indicate significant differences from baseline;
2) ##p<0.01 indicates significant difference from placebo group.

Compared to baseline, DR.ERGO^®^ significantly decreased visible spots (4.5% and 8.5%), latent spots (3.9% and 6.0%), brown spots (4.8% and 6.6%), and wrinkles (3.2% and 6.1%) (all p<0.05). In contrast, placebo significantly increased all counts (visible spots: 3.3% and 5.4%; latent spots: 4.9% and 7.4%; brown spots: 5.2% and 7.1%; wrinkles: 7.6% and 10.8%; all p<0.05).

#### 3.5.2. Qualitative Results (Scores)

As shown in Table 5-2, compared with the placebo group, the DR.ERGO^®^ group showed significant improvements in several skin parameters. Visible spot scores were significantly reduced at both week 4 (124.5%) and week 8 (478.9%) (p < 0.05). Brown spot scores decreased by 251.3% and 170.6% at weeks 4 and 8, respectively (p < 0.05), and latent spot scores decreased by 126.0% and 86.8% (p < 0.05). Wrinkle scores also showed decreasing trends at weeks 4 and 8 (126.6% and 195.4%, respectively; p < 0.1).

**Table 5-2.** Changes in Visible Spots, UV Spots, Brown Spots, and Wrinkles.

| Score | Time Point | Measured values |  | p-value<sup>2)</sup> |
| --- | --- | --- | --- | --- |
|  |  | DR.ERGO<sup>®</sup><br>Group (n=32)<sup>1)</sup> | Placebo Group<br>(n=34)<sup>1)</sup> |  |
| Visible Spots | a Before start | 26.653 ± 6.245 | 26.382 ± 7.553 |  |
|  | b After 4 weeks | 28.212 ± 6.842 | 26.230 ± 8.203 |  |
|  | Δa-b | 1.560 ± 3.169** | -0.152 ± 4.054 | 0.062<sup>‡</sup> |
|  | c After 8 weeks | 28.357 ± 6.936 | 25.932 ± 7.388 |  |
|  | Δa-c | 1.704 ± 3.269** | -0.450 ± 4.094 | 0.022<sup>#</sup> |
| Hidden Spots | a Before start | 25.667 ± 4.894 | 22.998 ± 5.553 |  |
|  | b After 4 weeks | 25.484 ± 4.858 | 23.701 ± 4.958 |  |
|  | Δa-b | -0.183 ± 3.918 | 0.703 ± 4.599 | 0.404 |
|  | c After 8 weeks | 25.805 ± 4.712 | 24.049 ± 4.837 |  |
|  |  | DR.ERGO<sup>®</sup><br>Group (n=32)<sup>1)</sup> | Placebo Group<br>(n=34)<sup>1)</sup> |  |
|  | Δa-c | 0.138 ± 4.671 | 1.051 ± 5.371 | 0.465 |
| Brown<br>Spots | a Before start | 22.441 ± 4.799 | 21.219 ± 4.843 |  |
|  | b After 4 weeks | 21.673 ± 4.803 | 21.727 ± 5.331 |  |
|  | Δa-b | -0.768 ± 4.865 | 0.508 ± 4.344 | 0.265 |
|  | c After 8 weeks | 21.947 ± 5.250 | 21.918 ± 5.431 |  |
|  | Δa-c | -0.494 ± 5.358 | 0.699 ± 4.487 | 0.329 |
| Wrinkles | a Before start | 24.458 ± 9.129 | 24.289 ± 8.848 |  |
|  | b After 4 weeks | 24.058 ± 8.031 | 25.790 ± 8.501 |  |
|  | Δa-b | -0.400 ± 3.120 | 1.501 ± 4.976<sup>†</sup> | 0.070<sup>‡</sup> |
|  | c After 8 weeks | 23.444 ± 7.966 | 25.352 ± 8.840 |  |
|  | Δa-c | -1.014 ± 3.345<sup>†</sup> | 1.063 ± 4.940 | 0.051<sup>‡</sup> |

Values are expressed as mean ± standard deviation.
1) <sup>†</sup>p<0.1, <sup>\*\*</sup>p<0.01 versus baseline (within-group comparison)
2) <sup>‡</sup>p<0.1, <sup>#</sup>p<0.05 versus placebo group (between-group comparison)

Relative to baseline, the DR.ERGO^®^ group exhibited significant reductions in visible spot scores at both week 4 and week 8 (p < 0.01). No significant changes were observed in latent spot, brown spot, or wrinkle scores, although wrinkle scores showed a trend toward reduction at week 8 (p < 0.1). In the placebo group, most parameters remained unchanged, except for a significant increase in latent spot scores at both time points (p < 0.01).

#### 3.5.3. Percentile Ranking Results

Table 5-3 presents the percentile changes. At weeks 4 and 8, wrinkle percentiles in the DR.ERGO^®^ group increased by 134.8% and 240.7%, respectively, though no significant differences were observed compared to the placebo group. In contrast, spot percentiles showed significant reductions of 654.9% and 2106.3% in the DR.ERGO^®^ group compared to placebo (p < 0.01).

**Table 5-3.** Percentile changes in Visible Spots, UV Spots, Brown Spots, and Wrinkles.

| Score | Time Point | Measured values |  | p-value<sup>2)</sup> |
| --- | --- | --- | --- | --- |
|  |  | DR.ERGO<sup>®</sup><br>Group (n=32)<sup>1)</sup> | Placebo Group<br>(n=34)<sup>1)</sup> |  |
| Visible<br>Spots | a Before start | 87.1 ± 9.2 | 84.5 ± 17.8 |  |
|  | b After 4 weeks | 83.5 ± 11.4 | 85.1 ± 16.8 |  |
|  | Δa-b | -3.7 ± 5.0<sup>**</sup> | 0.7 ± 7.0 | 0.005<sup>##</sup> |
|  | c After 8 weeks | 82.1 ± 13.5 | 84.7 ± 16.6 |  |
|  | Δa-c | -5.0 ± 6.8<sup>**</sup> | 0.3 ± 7.4 | 0.004<sup>##</sup> |
| Hidden<br>Spots | a Before start | 66.0 ± 15.5 | 60.9 ± 20.2 |  |
|  | b After 4 weeks | 51.9 ± 19.1 | 46.2 ± 21.3 |  |
|  | Δa-b | -14.1 ± 15.8<sup>**</sup> | -14.7 ± 12.0<sup>**</sup> | 0.869 |
|  | c After 8 weeks | 46.4 ± 19.0 | 42.7 ± 19.9 |  |
|  |  | DR.ERGO<sup>®</sup><br>Group (n=32)<sup>1)</sup> | Placebo Group<br>(n=34)<sup>1)</sup> |  |
| | Δa-c | -19.6 ± 15.2** | -18.2 ± 11.0** | 0.675 |
| Brown<br>Spots | a Before start | 46.8 ± 18.0 | 45.1 ± 23.5 | |
| | b After 4 weeks | 45.1 ± 17.0 | 42.2 ± 19.6 | |
| | Δa-b | -1.7 ± 14.9 | -2.9 ± 14.3 | 0.740 |
| | c After 8 weeks | 42.3 ± 17.3 | 41.4 ± 18.1 | |
| | Δa-c | -4.4 ± 17.5 | -3.7 ± 15.1 | 0.862 |
| Wrinkles | a Before start | 73.8 ± 23.0 | 73.8 ± 24.0 | |
| | b After 4 weeks | 74.4 ± 19.7 | 72.0 ± 24.5 | |
| | Δa-b | 0.6 ± 9.1 | -1.8 ± 12.6 | 0.376 |
| | c After 8 weeks | 76.8 ± 17.2 | 71.7 ± 23.9 | |
| | Δa-c | 3.0 ± 11.4 | -2.1 ± 12.7 | 0.090<sup>‡</sup> |

Values are expressed as mean ± standard deviation.
1) \*\*p<0.01 versus baseline (within-group comparison)
2) ‡p<0.1, ##p<0.01 versus placebo group (between-group comparison)

Longitudinally, wrinkle percentiles in the DR.ERGO^®^ group showed numerical increases at weeks 4 and 8 (0.8% and 4.1%), but these changes were not statistically significant. Latent spot percentiles were significantly reduced in both the DR.ERGO^®^ group (21.4% and 29.7% decrease; p < 0.01) and the placebo group (24.1% and 29.9% decrease; p < 0.01). No statistically significant changes were observed in brown spot percentiles in either group.

Representative VISIA photographs of participants at baseline (Week 0), Week 4, and Week 8 illustrate visible improvements in pigmentation appearance in the ergothioneine group (Figure 2). Notable reductions in visible spots, latent spots, and brown spots were observed in alignment with instrumental measurements.

**Figure 2.**
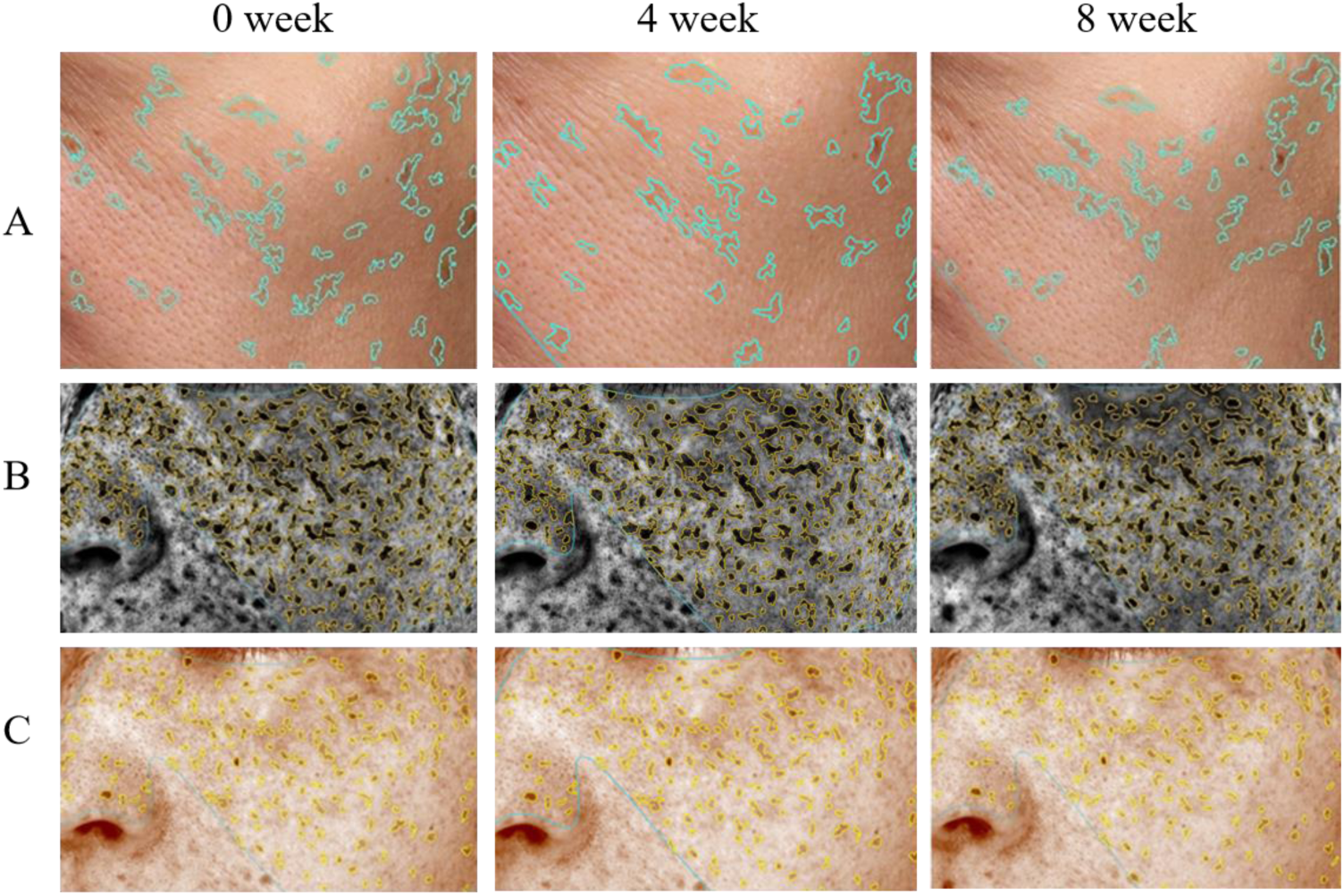
Representative VISIA images of a participant in the test group at baseline (Week 0), Week 4, and Week 8. Improvements in visible spots (A), latent spots (B), and brown spots (C) are observed over time.

### 3.6. Safety Evaluation

Based on the results of physical assessments and adverse event monitoring through participant logs, no adverse events were reported during the study. Furthermore, no clinically relevant side effects attributable to the test product were observed.

## 4. Discussion

This randomized, double-blind, placebo-controlled clinical trial demonstrated that daily oral supplementation with 30 mg of ergothioneine over an 8-week period significantly improved multiple parameters of skin condition in healthy adult women with perceived skin aging. The ergothioneine group exhibited statistically greater reductions in melanin and erythema indices, increases in skin gloss and elasticity (R2, R5, R7), and decreases in visible spots, latent spots, brown spots, and wrinkles compared to placebo.

### 4.1. Interpretation of Key Findings

The improvement in melanin and erythema indices suggests that ergothioneine may reduce hyperpigmentation and local inflammation—hallmarks of photoaging and oxidative stress-induced skin damage. Skin glossiness, which reflects the skin’s ability to reflect light, was also significantly enhanced, indicating improved surface smoothness and moisture retention.

Elasticity is a critical determinant of youthful skin, and ergothioneine significantly improved all three key elasticity metrics (R2: gross elasticity, R5: net elasticity, and R7: biological elasticity). These findings suggest that ergothioneine may support dermal matrix integrity, possibly by preserving collagen and elastin or reducing glycation-induced rigidity.

Notably, the reduction in pigmentation spots and wrinkles was supported by instrumental VISIA analysis, which showed both numerical and percentile improvements. These results were consistent across weeks 4 and 8, indicating both early onset and sustained efficacy.

### 4.2. Comparison with Existing Literature

Previous in vitro and in vivo studies have identified ergothioneine as a potent antioxidant capable of scavenging reactive oxygen and nitrogen species, protecting mitochondrial DNA, and modulating inflammatory pathways such as NF-κB and Nrf2 (Chen et al., 2024; H. M. Liu et al., 2023). Studies in human dermal fibroblasts have shown that ergothioneine protects against UVA-induced oxidative damage, decreases expression of matrix metalloproteinases (MMPs), and enhances collagen synthesis (Hseu et al., 2015; Hseu et al., 2020; Y. Li et al., 2024). However, clinical studies on its dermatological effects remain scarce.

These findings corroborate and extend previous clinical evidence from (Hanayama et al., 2024), who investigated an ergothioneine-rich mushroom extract (Pleurotus sp., 25 mg ergothioneine/day) in a 12-week randomized double-blind trial, and (Chunyue Zhang, 2023), who examined pure ergothioneine supplementation (25 mg/day) in a 4-week open-label study. Below, we contextualize our results within this existing literature by comparing key outcomes.

#### 4.2.1. Skin Hydration and Barrier Function

Our study did not directly assess skin hydration, whereas Hanayama et al. (2024) reported a significant increase in skin moisture content (temple: +5.8%, p < 0.05; arm: +9.0%, p < 0.05) and reduced transepidermal water loss (TEWL) after 12 weeks of mushroom-derived ergothioneine supplementation. These improvements correlated with elevated plasma ergothioneine levels (r = 0.168, p = 0.038), suggesting systemic absorption mediates hydration benefits. Zhang et al. (2023) indirectly supported these findings by showing reduced pore size and porphyrins (indicators of sebum balance) after 4 weeks of pure ergothioneine supplementation. While our trial focused on elasticity and pigmentation, the consistency across studies implies ergothioneine enhances skin hydration through both direct (e.g., antioxidant effects) and indirect (e.g., sebum regulation) mechanisms.

#### 4.2.2. Skin Elasticity

A novel contribution of our study is the quantification of ergothioneine’s effects on biomechanical elasticity parameters (R2, R5, R7), which were not evaluated in Zhang et al. (2023) or Hanayama et al. (2024). We observed significant improvements in gross (R2: +4.7%, p < 0.01) and biological elasticity (R7: +8.4%, p < 0.01) after 8 weeks, contrasting with placebo declines. These results align with preclinical evidence that ergothioneine mitigates collagen degradation by suppressing MMP-1 and enhancing Nrf2-mediated antioxidant pathways (Hseu et al., 2020). The absence of elasticity data in prior trials highlights a gap our study addresses.

#### 4.2.3. Pigmentation and Wrinkles

All three studies reported anti-pigmentary and anti-wrinkle effects, though metrics differed.

Pigmentation: Zhang et al. (2023) noted reductions in UV spots (+18.4%, p < 0.01) and brown spots (+16.6%, p < 0.01) via VISIA, while our trial showed decreases in melanin index (−4.5%, p < 0.05) and erythema (−5.4%, p < 0.05). Hanayama et al. (2024) did not detect significant changes in UV spots but reported improved texture scores (p < 0.05).

Wrinkles: Our study and Zhang et al. (2023) observed early improvements (4-8 weeks), with wrinkle counts declining by 6.1% (p < 0.05) and VISIA scores increasing by 21.6% (p < 0.01), respectively. Hanayama et al. (2024) demonstrated effects emerging later (12 weeks), suggesting duration-dependent efficacy.

These variations may reflect differences in ergothioneine formulation (pure vs. mushroom-derived), dosage (25-30 mg/day), or participant demographics (age, baseline skin condition).

#### 4.2.4. Safety and Tolerability

Consistent across studies, ergothioneine was well tolerated at 25-30 mg/day over 4-12 weeks, with no adverse events reported. Plasma ergothioneine levels in Hanayama et al. (2024) (15.9 μM) and our trial (dose-dependent increase) confirm bioavailability, supporting its use as a nutraceutical.

### 4.3. Mechanistic Insights into Ergothioneine’s Dermatological Effects

Recent advances in mechanistic research have substantially deepened our understanding of how orally administered ergothioneine (ergothioneine) may exert dermatological benefits at the cellular and molecular levels. Central to its bioactivity is the highly selective OCTN1 (SLC22A4) transporter, which facilitates efficient uptake and mitochondrial accumulation of ergothioneine in oxidative stress-prone tissues such as the skin. Notably, OCTN1 is preferentially expressed in basal and granular epidermal layers, where cellular renewal and barrier maintenance are most active. Once internalized, ergothioneine localizes to mitochondria, where it directly scavenges reactive oxygen species (ROS) and protects mitochondrial DNA from UV- and inflammation-induced damage (Cheah & Halliwell, 2012; Yongchao Li et al., 2024; Y. Li et al., 2024).

At the signaling level, ergothioneine activates key protective pathways such as the Nrf2/ARE axis, enhancing the expression of antioxidant enzymes including HO-1, NQO1, and γ-GCLC. These enzymes contribute to redox homeostasis and glutathione regeneration, reinforcing cellular defense systems against photoaging and environmental insult (Hseu et al., 2015; Hseu et al., 2020; Y. Li et al., 2024). In parallel, ergothioneine modulates the PI3K/Akt/Nrf2 and SIRT1/Nrf2 pathways, which are implicated in collagen preservation, inflammation resolution, and mitochondrial maintenance. These pathways converge to reduce matrix metalloproteinase (MMP) activity, enhance collagen synthesis, and suppress pro-inflammatory cytokines (TNF-α, IL-6, IL-1β), all of which are central to maintaining skin structure and function during aging (Chen et al., 2023; Yongchao Li et al., 2024; Paul, 2022).

Additional mechanisms include AP-1 pathway inhibition, which further contributes to reduced MMP-1 expression and prevention of collagen degradation, as well as non-competitive inhibition of tyrosinase, potentially mitigating hyperpigmentation and skin tone unevenness (Hseu et al., 2020; H.-M. Liu et al., 2023). Moreover, ergothioneine has recently been shown to protect against blue light-induced oxidative stress, a modern environmental factor increasingly associated with premature aging. Through its integration into the endogenous antioxidant network, ergothioneine also enhances the activity of vitamins C and E and supports glutathione and NADPH cycling, indicating a synergistic role in cutaneous antioxidant defense (Cheah & Halliwell, 2012; Y. Li et al., 2024).

Together, these findings establish a multi-layered mechanistic foundation by which oral ergothioneine may protect, repair, and rejuvenate the skin at both cellular and systemic levels. Such mechanistic evidence strongly complements the clinical outcomes observed in this trial and reinforces the therapeutic potential of ergothioneine as a scientifically validated nutricosmetic agent.

### 4.4. Clinical Significance

The trial population—healthy women aged 35 to 59 years reporting signs of skin aging—represents a relevant consumer segment in the global nutricosmetic market. The high compliance rate (94.3%) and absence of adverse events underscore the practicality and safety of ergothioneine supplementation in this demographic. The consistent improvements suggest meaningful benefit that may translate into high consumer satisfaction and long-term use.

Given its excellent safety profile, ergothioneine may be particularly valuable in populations seeking non-invasive, daily skincare regimens. The early improvements observed at 4 weeks also enhance its commercial appeal as a fast-acting supplement.

This trial represents one of the most comprehensive investigations to date evaluating pure ergothioneine’s effects on skin aging through both instrumental and perceptual endpoints

### 4.5. Strengths, Limitations, and Future Directions

This study has several methodological strengths. It employed a randomized, double-blind, placebo-controlled design, enhancing internal validity and minimizing bias. The integration of validated instrumental assessments—including melanin and erythema indices, skin gloss, elasticity parameters, and VISIA-based wrinkle and spot analyses—provided robust, objective measures of skin health. High participant retention and adherence further support the reliability of the findings.

Despite these strengths, several limitations should be acknowledged. First, the study cohort consisted solely of Japanese women aged 35–59 years, which may limit generalizability across sexes, ethnicities, and age groups. Second, the 8-week intervention period, while sufficient to detect short-term effects, does not allow conclusions about the sustainability of benefits or the risk of relapse upon discontinuation. Third, the placebo group also showed modest improvements in self-perception—highlighting the well-documented placebo response in beauty and wellness studies. Moreover, this study focused on a single daily dosage (30 mg/day) without evaluating dose–response relationships, and hydration-specific endpoints such as corneometry or transepidermal water loss (TEWL) were not included.

Future research should build upon these findings by conducting multicenter, longer-duration clinical trials involving more diverse populations to assess cross-ethnic and age-related efficacy. Extended follow-up periods (≥12 weeks) will help determine whether the observed benefits are sustained and whether continued supplementation is necessary. Mechanistic studies should be designed to directly assess ergothioneine’s influence on skin biomarkers—such as collagen content, inflammatory cytokines, and oxidative stress markers—to better understand the biological underpinnings of its effects. Comparative trials with other nutricosmetic agents or topical skincare regimens are warranted to position ergothioneine within the broader anti-aging landscape. Finally, exploring synergistic combinations with other bioactive compounds (e.g., vitamin C, hyaluronic acid, glutathione) may further enhance its clinical utility as part of an integrative nutricosmetic strategy.

Together, these directions can help refine the therapeutic positioning of oral ergothioneine and unlock its full potential as a next-generation ingredient in systemic skin health and anti-aging interventions.

## 5. Conclusion

This 8-week, randomized, double-blind, placebo-controlled clinical trial evaluated the effects of daily oral intake of an ergothioneine-containing lozenge on skin elasticity and overall skin condition in women aged 35 to 59 years who reported signs of skin aging. Participants were randomly assigned to receive either the test product or a placebo. After 8 weeks of supplementation, the test group showed statistically significant improvements across multiple skin parameters compared to the placebo group, including a* value (redness), melanin index, erythema, skin radiance, elasticity metrics (R2, R5, R7), number of visible spots, latent spots, brown spots, and wrinkle count. Additionally, no adverse events or side effects were reported, indicating that the test product was safe and well tolerated under the conditions of this trial.

## Data Availability

All data produced in the present study are available upon reasonable request to the authors.
All data produced in the present work are contained in the manuscript.

## Acknowledgements

The authors wish to acknowledge Shanghai ergothioneine SYNBIO Group Co., Ltd., for providing the test supplement and Japan Clinical Trial Association (JACTA) for the clinical experimentation.

